# Does Gender or Religion Contribute to the Risk of COVID-19 in Hospital Doctors in the UK?

**DOI:** 10.1101/2020.06.15.20125450

**Authors:** Sunil Daga, Sadaf Jafferbhoy, Geeta Menon, Mansoor Ali, Subarna Chakravorty, Saqib Ghani, Amir Burney, JS Bamrah, Ramesh Mehta, Indranil Chakravorty

## Abstract

The novel coronavirus pandemic is posing significant challenges to healthcare workers (HCWs) in adjusting to redeployed clinical settings and enhanced risk to their own health. Studies suggest a variable impact of COVID-19 based on factors such as age, gender, comorbidities and ethnicity. Workplace measures such as personal protective equipment (PPE), social distancing (SD) and avoidance of exposure for the vulnerable, mitigate this risk. This online questionnaire-based study explored the impact of gender and religion in addition to workplace measures associated with risk of COVID-19 in hospital doctors in acute and mental health institutions in the UK.

The survey had 1206 responses, majority (94%) from BAME backgrounds. A quarter of the respondents had either confirmed or suspected COVID-19, a similar proportion reported inadequate PPE and 2/3 could not comply with SD. One third reported being reprimanded in relation to PPE or avoidance of risk. In univariate analysis, age over 50 years, being female, Muslim and inability to avoid exposure in the workplace was associated with risk of COVID-19. On multivariate analysis, inadequate PPE remained an independent predictor with a twofold (OR 2.29, (CI - 1.22-4.33), p=0.01) risk of COVID-19.

This study demonstrates that PPE, SD and workplace measures to mitigate risk remain important for reducing risk of COVID-19 in hospital doctors. Gender and religion did not appear to be independent determinants. It is imperative that employers consolidate risk reduction measures and foster a culture of safety to encourage employees to voice any safety concerns. (240 words)

## Introduction

The COVID-19 pandemic has posed a global threat affecting people from all backgrounds (1). Healthcare workers (HCWs) inevitably carry a high risk of contracting the disease (2,3). Several studies have shown significant disparity in the severity of COVID-19 and outcomes based on ethnicity, among other factors (4-8). Multiple factors including comorbidities and social deprivation have been proposed to contribute to high mortality in Black, Asian and Minority Ethnic (BAME) people (5-8). Even after adjusting for inherent differences, people of BAME backgrounds are twice as likely to die from COVID-19 as compared to their white counterparts (9). This is also seen in HCWs in the UK National Health Service (NHS), where the BAME community makes up 20% of the overall workforce but accounts for two-thirds of COVID-19 related deaths (10). Furthermore, BAME doctors form 44% of NHS doctors, and 94% of the mortality statistics (11).

Population based data from China and Italy has shown that men appear to be at a higher risk of COVID-19 infection (12,13). However, studies from HCWs in other countries suggest a higher proportion of females (average 70%) in COVID-19 (14,15). This may be due to a higher number of frontline HCWs being female. At least one analysis of HCW who died in the UK showed that 39% of nurses and 94% of doctors were male. In addition, among those who died, 71% of nurses and 94% of doctors were from BAME backgrounds. (11). Gender differences have been observed in other outbreaks such as Severe Acute Respiratory Syndrome and Middle Eastern Respiratory Syndrome, where significantly higher fatality rates were reported in males (16,17). The disproportionately high death rate from COVID-19 in HCWs from BAME background appears to be only partially explained by age, gender, socio-demographic features and underlying health conditions (6,8).

Thirteen percent of respondents in the NHS Staff survey in 2019 reported discrimination; due to ethnicity (45%), gender (22%) and religion (6%) (18,19). The same survey showed 31% staff experienced bullying or harassment at work. A survey conducted by the British Medical Association (BMA) during this pandemic suggested 64% of staff from BAME background felt pressured to work in settings with inadequate personal protective equipment (PPE) as compared to 33% of their white colleagues (20,21). Studies from our group have previously shown that, amongst HCWs, BAME background and adverse workplace measures were predictors of higher risk of COVID-19 (22,23).

A meta-analysis has confirmed that viral spread is reduced with the use of eye protection, face masks and social distancing (of greater than one metre) in healthcare settings and supports their use in minimising exposure to healthcare staff (24). Various risk assessment frameworks and scores now include ethnicity and gender as variables (25,26,27), however these appear to be mostly based on extrapolation of data obtained from population studies, rather than specific data on HCWs. This study explores the contribution of gender and religious identity in addition to workplace measures, as well as being reprimanded (for asking or wearing PPE) in risk analysis for hospital doctors who have self-reported COVID19.

### Aims

An online survey was designed to explore the hypothesis that hospital doctors had a variable risk of COVID-19, due to differential treatment based on their gender or religion.

This would manifest in differential rates of (a) access to PPE, (b) compliance with social distancing (SD) at work and (c) access to employer supported self-isolation (SI) when identified as ‘vulnerable’ based on Public Health England (PHE) guidance (28,29).

Primary outcome was a self-reported diagnosis of COVID-19 confirmed by a positive viral swab test or self-isolation with symptoms of COVID-19 as per PHE guidance where a test was not undertaken.

### Method

An anonymous, online <u>survey</u> using Survey Monkey® was undertaken which was open to hospital doctors from acute and mental health NHS Trusts in the United Kingdom. The survey was designed and distributed by British Association of Physicians of Indian Origin (BAPIO) Institute for Health Research (BIHR) & Education subcommittee of the Association of Pakistani Physicians of Northern Europe (APPNE). The survey questions are available in the appendix. Data variables collected are shown in Table 1. The study was reviewed by institutional review board, BIHR; and no formal ethics review was required to conduct this survey.

**Table 1:** Data variables used to collect the responses for the survey.

| **Variables** |
| --- |
| **Demographics**<br>a) Clinical setting - Teaching hospital, non-teaching hospital or Mental health trust<br>b) Gender - Male, Female, Transgender, Prefer not to disclose<br>c) Number of additional family members in the same household<br>d) Ethnicity - Caucasian (British/Irish Traveller/Any); South Asian- Indian /Pakistani/Bangladeshi/Other; Black/ African/ Afro-Caribbean/ African-American; Arab/ Middle-eastern/ North African; Chinese/ SE Asian ; Mixed; Any other ethnicity and Do not wish to declare)<br>e) Religion - Christian, Muslim, Hindu, Buddhist, Jewish, Sikh, No religion, Do not want to state, Other<br>f) Age groups (20-30;30-40; 40-50; 50-60; 60-70;70+ years)<br>g) Any comorbidities - Highly Vulnerable (where you have been asked to shield and stay at Home); Vulnerable (where social distancing at work and home/community is recommended) or Healthy and none of above. |
| **Workplace measures**<br>a) Work in areas - Patients with COVID-19 (suspected or confirmed) are cared for; Patients with Non-COVID-19 only are cared for; Both<br>b) Access to PPE to do the job safely - Strongly agree; Somewhat agree; Neither agree nor disagree ; Somewhat disagree; Strongly disagree<br>c) Able to comply SD at work - All of the times; Most of the times; Some of the times; A few of the times; None of the times<br>d) Able to negotiate changes in work - Work from home; Work in low COVID-19 risk areas; Virtual consultations; None allowed by employer; Other; Not applicable<br>e) Reprimanded from wearing or asking PPE - Always; Usually; Sometimes; Rarely; Never<br>f) Redeployed to an area that cares for - COVID-19 patients (Suspected or confirmed); Non-COVID-19 patients; Both of the above; None of the above |
| **COVID-19 status**<br>• Never had suspected or confirmed COVID-19 infection<br>• Confirmed COVID-19 infection (by a PCR test)<br>• Suspected COVID-19 infection<br>• Self-isolating due to exposure to suspected or confirmed case of COVID-19 infection at Home<br>• None of above options |

The online survey link was sent to all members of both the organisations and doctors from wider communities in the UK, using email and social media. The survey specified an implied consent to share the data and results with appropriate agencies or organisations involved directly or indirectly in HCWs and COVID-19 pandemic measures. No personal identifiable information was collected. Data was stored at the BAPIO/BIHR office in compliance with UK General Data Protection Regulations.

### Study population & Statistical Analysis

A convenience sample of survey responses was planned to be collected over a four-week period; similar to previous surveys (22,23). The survey results are reported as cross-tabulation of proportions between the different primary categorical variables (based on gender and religion). Descriptive statistics were used for primary categorical variables. Univariate analysis was conducted between groups of categorical variables using Fisher Exact 2-tailed test (GraphPadPRISM®). Multivariable model was constructed including demographic (age-group and number of household members, ethnicity, gender, religions), clinical setting (teaching, non-teaching or mental health trusts) and exposure to COVID-19 (areas caring for COVID-19 patients, PPE, SD). Non-significant variables from univariate analysis were excluded from final models. Regression analysis was conducted using SPSS v26 software (IBM Inc., USA) and reported as odds ratio (OD), 95% confidence interval (CI) and significant when p-value <0.05 (non-significant values were reported as ‘ns’).

## Results

### (i) Population

The survey received 1206 responses between 26 April and 29 May 2020. Table 2 shows the characteristics of the respondents and Table 3 summarises the status of workplace measures reported by the respondents. Majority (65.6%) were working in a teaching hospital setting, 38.8% were over 50 years of age, 70.9% were male, 93.7% were from BAME background and their religious identities were Hindu (44.5%), Muslim (32.1%) or Christian (10.4%).

**Table 2:** Characteristics of respondents to survey (*as per PHE (28,29))

| Variables | All |
| --- | --- |
| Number | 1206 |
| Institution |  |
| Teaching | 791 (65.59%) |
| Non-teaching | 266 (22.06%) |
| Mental Health | 149 (12.35%) |
| Household members |  |
| ≤ 5 | 1165 (96.60%) |
| >5 | 41 (3.40%) |
| Ethnicity |  |
| BAME | 1130 (93.70%) |
| White | 72 (6%) |
| Did not wish to state | 4 (0.33%) |
| Age |  |
| ≤ 50 | 738 (61.19%) |
| >50 | 468 (38.81%) |
| Gender |  |
| Male | 855 (70.9%) |
| Female | 347 (28.8%) |
| Prefer not to state | 4 (0.33%) |
| Religion |  |
| Christian | 125 (10.37%) |
| Hindu | 537 (44.53%) |
| Muslim | 387 (32.1%) |
| Other religion | 45 (3.73%) |
| Did not wish to state | 40 (3.32%) |
| No religion | 72 (5.97%) |
| Health status* |  |
| Vulnerable | 303 (25.12%) |
| Healthy | 903 (74.88%) |

**Table 3:** Workplace measures - overall and distribution as per religion and gender. *(responses from Christian religion - expressed as all and separate BAME-Christian to allow comparison of proportions with responses from Muslim and Hindu religion which were all from BAME ethnicity)*

| Variables | All | All-Christian | BAME-Christian | Muslim | Hindu | Female | Male |
| --- | --- | --- | --- | --- | --- | --- | --- |
| **Number** | 1206 | 125 | 95 | 387 | 537 | 347 | 855 |
| **Ward area** |  |  |  |  |  |  |  |
| Non-COVID-19 Only | 151 (12.5%) | 12 (9.6%) | 12 (12.6%) | 52 (13.4%) | 67 (12.5%) | 48(13.8%) | 103 (12.05) |
| COVID-19 | 1055 (87.5%) | 113 (90.4%) | 83 (87.4%) | 335 (86.6%) | 470 (87.5%) | 299 (86.2%) | 752 (87.95) |
| **Access to PPE** |  |  |  |  |  |  |  |
| Agree | 739 (61.3%) | 77 (61.6%) | 55(57.9%) | 217 (56.1%) | 348 (64.8%) | 208 (60%) | 529 (61.9%) |
| Disagree | 315 (26.1%) | 28 (22.4%) | 26 (27%) | 118 (30.5%) | 129 (24%) | 94 (27%) | 220 (25.7%) |
| Neither agree or disagree | 152 (12.6%) | 20 (16%) | 14 (14.7%) | 52 (13.4%) | 60 (11.2%) | 45 (13%) | 106 (12.4%) |
| **Able to comply with Social Distancing** |  |  |  |  |  |  |  |
| Most/all | 430 (35.7%) | 46 (36.8%) | 36 (37.9%) | 139 (35.9%) | 193 (36%) | 120 (34.6%) | 308 (36%) |
| Some, few or none | 776 (64.3%) | 79 (63.2%) | 59 (62%) | 248 (64.1%) | 344 (64%) | 227 (65.4%) | 547 (64%) |
| **Able to negotiate** |  |  |  |  |  |  |  |
| None | 224 (18.6%) | 18 (14.4%) | 17 (17.9%) | 69 (17.8%) | 105 (19.6%) | 57(16.4%) | 166 (19.4%) |
| Yes | 831 (68.9%) | 85 (68%) | 45 (47%) | 271 (70%) | 378 (70.4%) | 222 (64%) | 607 (71%) |
| Not applicable | 151 (12.5%) | 22 (17.6%) | 33 (34.7%) | 47 (12.1%) | 54 (10%) | 68 (19.6%) | 82 (9.6%) |
| **Reprimanded for PPE** |  |  |  |  |  |  |  |
| yes | 360 (29.9%) | 28 (22.4%) | 24 (25.3%) | 135 (34.9%) | 154 (28.7%) | 98 (28%) | 259 (30%) |
| Rare/Never | 846 (70.1%) | 97 (77.6%) | 81 (85.3%) | 252 (65.1%) | 383 (71.3%) | 249 (72%) | 596 (70%) |
| **Redeployed to area that cares** |  |  |  |  |  |  |  |
| Non-COVID-19 only | 68 (5.6%) | 4 (3.2%) | 3 (3.2%) | 32 (8.3%) | 27 (5%) | 13 (3.7%) | 55 (6.4%) |
| COVID-19 | 455 (37.7%) | 41 (32.8%) | 27 (28.4%) | 164 (42.4%) | 200 (37.3%) | 130 (37.5%) | 323 (37.8%) |
| Not applicable | 683 (56.6%) | 80 (64%) | 65 (68.4%) | 191 (49.4%) | 310 (57.7%) | 204 (58.8%) | 477 (55.8%) |

About a quarter identified themselves as ‘vulnerable’ according to PHE defined criteria. Age distribution of the respondents is shown in Figure 1. COVID-19 diagnosis was confirmed in 104 (8.6%) and, 213 (17.7%) were in self-isolation due to symptoms compatible with COVID-19 (suspected COVID-19) as shown in Figure 2.

**Figure 1:**
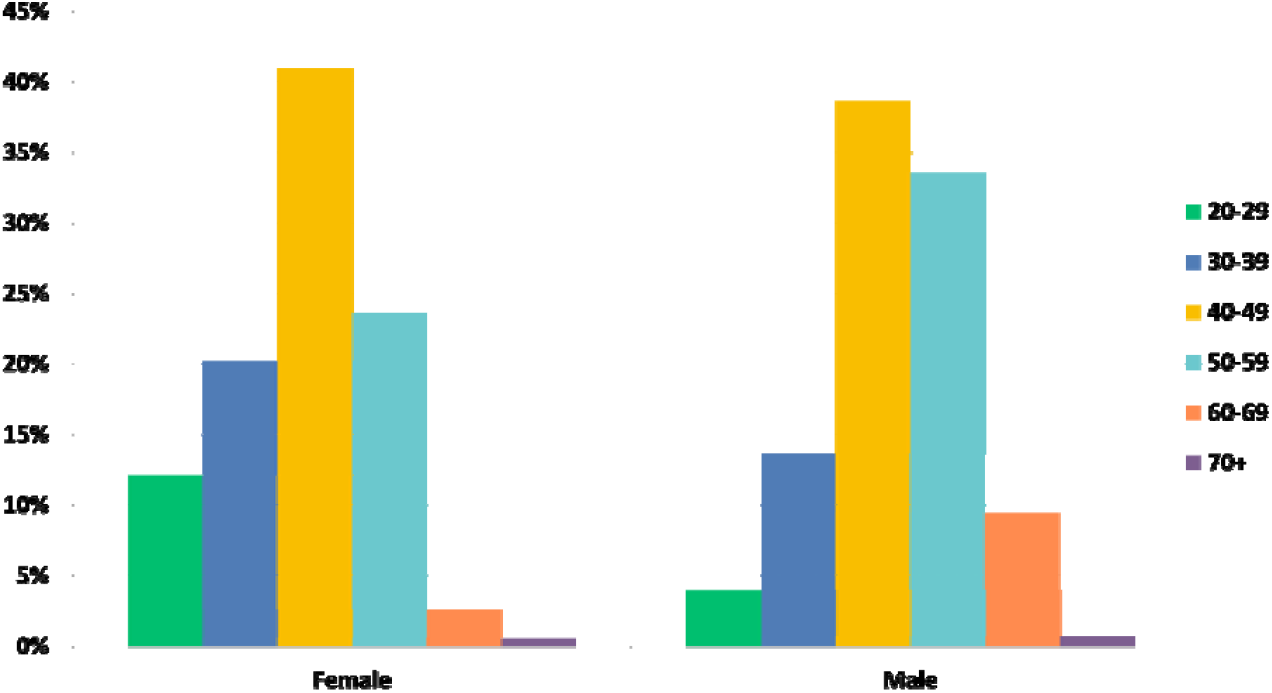
Age distribution as per gender of the respondents.

**Figure 2:**
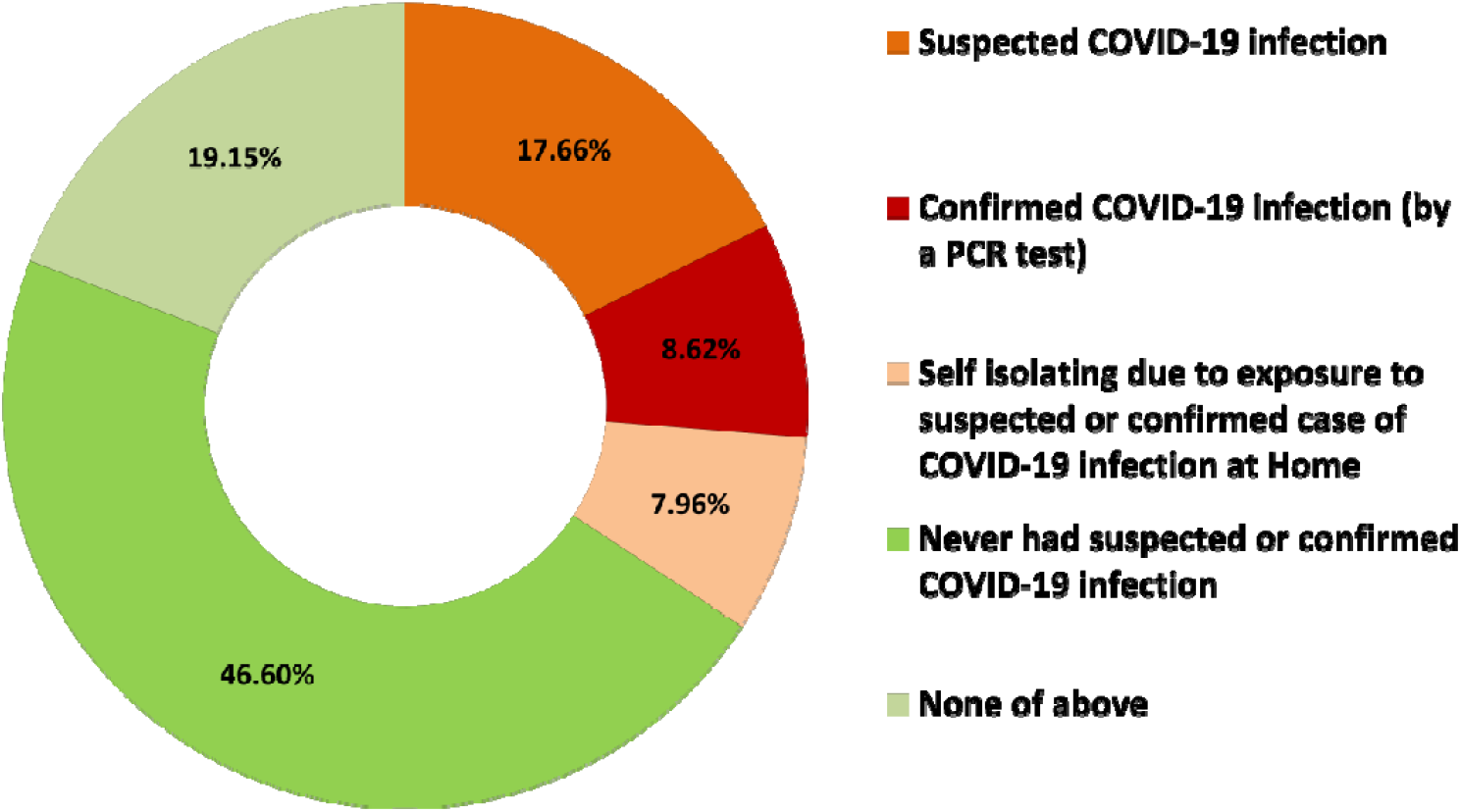
COVID-19 related status of the respondents.

Workplace measures are summarised in Table 3, Figures 3 and 4. 87.5% of respondents who reported COVID-19 were working in areas that cared for patients with suspected or confirmed COVID-19. Only 61.3% reported (strongly or somewhat) adequate access to PPE, 35.7% were able to comply with SD (most or all times) and 68.9% were able to negotiate different working environment to reduce risk. There was an incremental rise in COVID-19 with inadequate PPE and inability to comply with SD at work (Figures 3 and 4).

**Figure 3:**
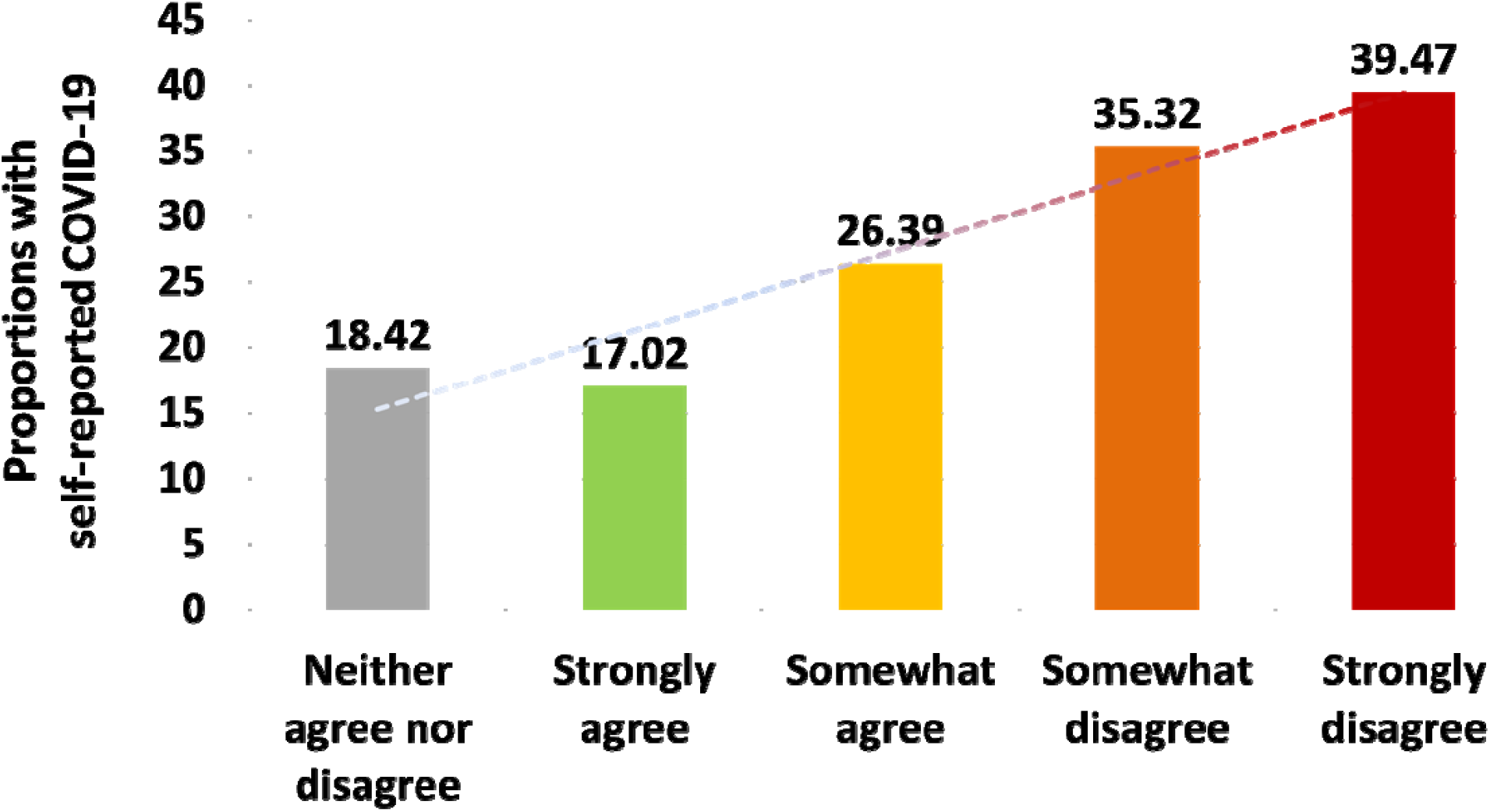
Proportions of self-reported COVID-19 in relation to adequate PPE.

**Figure 4:**
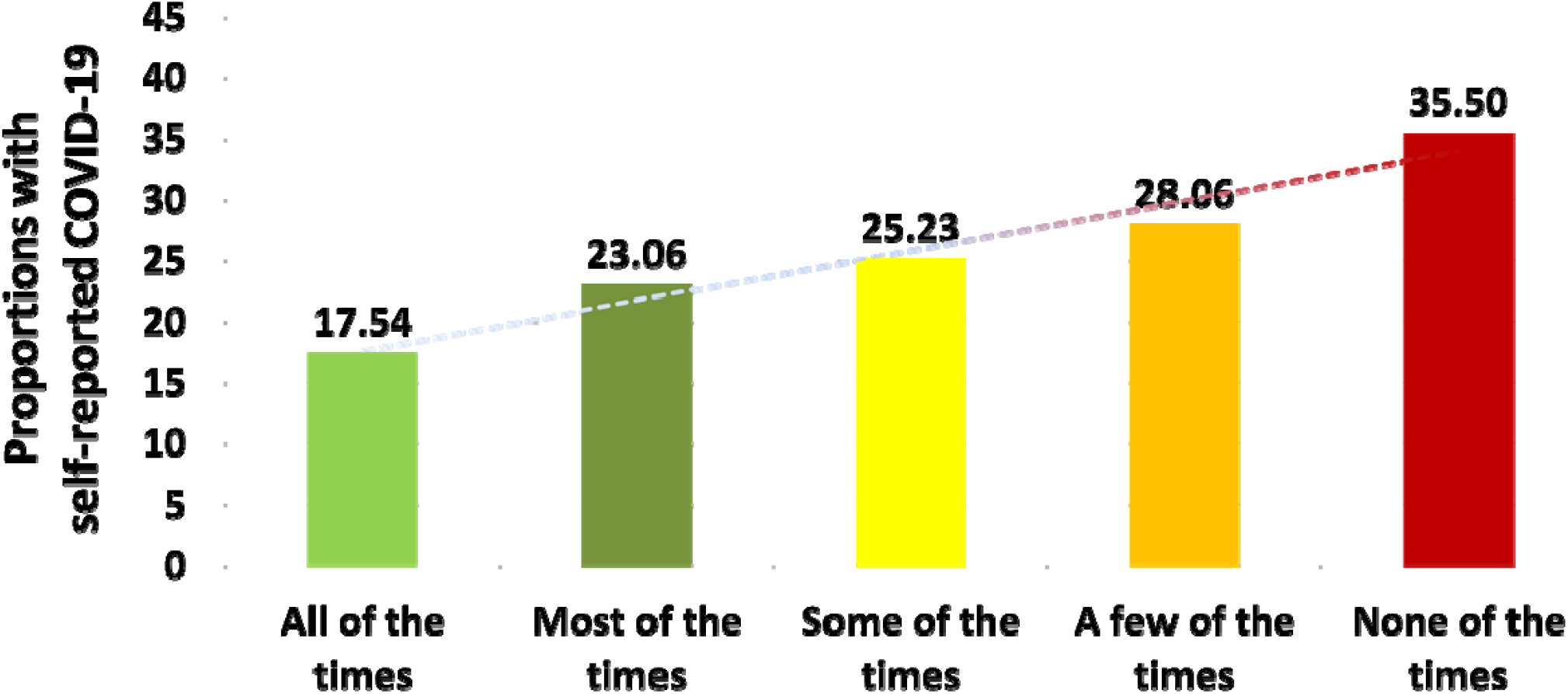
Proportions of self-reported COVID-19 in relation to ability to comply with SD.

### (ii) The Impact of Gender

In our survey, 1202 responses were included for analysis to explore gender differences in risk of COVID-19. Female respondents were of a younger age (73.2 % versus 56.3 %; p <0.0001) and a lower proportion identified as ‘vulnerable’.

Our analysis showed no gender differences in accessing to PPE, ability to comply with SD, redeployment or working in high risk areas (Table 3). A higher proportion of female respondents reported confirmed or suspected COVID-19 (30% versus 25%; p =0.04). Male respondents had a higher proportion of confirmed cases (9.6% versus 6.3%, p =0.07).

### (iii) The Impact of Religion

The majority of respondents identified themselves as Christians (n=125, 10.4 %), Muslim (n=387, 32.1%) or Hindu (n=537, 44.5%). The remaining respondents (n = 157, 13%) who identified themselves as Sikh, Jewish, Buddhist, or with no religion (see Table 3) were in small numbers and thus were excluded from analysis. Amongst Christians, 95 were from BAME background, whilst 30 were white.

Access to PPE was significantly lower (24% versus 30.5%, p = 0.0133) amongst Muslims. A higher proportion of Muslims (35 % versus 28%, p =0.01) reported being reprimanded for wearing or asking for PPE. There was no difference between Hindu or Muslim respondents in other workplace variables, such as working with COVID-19 patients, ability to practice SD or redeployment compared to either overall Christian or BAME-Christian respondents.

In this cohort, Muslims had a higher prevalence of suspected or confirmed COVID-19 compared to Hindus (34.6 % versus 19.3%, OR 2.18; 95% CI 1.52 - 2.95, p <0.001); but not to ‘all Christians (29.6%)’ or ‘BAME-Christians (28.4%)’.

### (iv) Risk of COVID-19

#### Univariate analysis

All variables related to workplace measures (except re-deployment) were significantly associated with higher risk of COVID-19 (Table 4). There was a higher self-reported COVID-19 in respondents below the age of 50 years. Other variables; such as number of household members, ethnicity or vulnerability were not significant (Table 4).

**Table 4:** Univariate analysis looking at risk factors for COVID19 (* Fisher Exact 2-tail test)

| Variables | COVID19 susp/+ve | COVID -ve | p value* |
| --- | --- | --- | --- |
| **Number** | 317 (26.28%) | 889 (72.71%) |  |
| **Institution** |  |  |  |
| Teaching | 210 (66.24%) | 581 (65.35%) | ref |
| Non-teaching | 69 (21.77%) | 197 (22.16%) | ns |
| Mental Health | 38 (11.99%) | 111 (12.49%) | ns |
| **Household members** |  |  | ns |
| ≤ 5 | 303 (95.58%) | 862 (96.96%) |  |
| >5 | 14 (4.41%) | 27 (3.04%) |  |
| **Ethnicity** |  |  | ns |
| BAME | 291 (91.80%) | 839 (94.40%) |  |
| White | 22 (6.94 %%) | 50 (5.60%) |  |
| **Age** |  |  | **0.0031** |
| ≤ 50 | 216 (68.14%) | 522 (58.72%) |  |
| >50 | 101 (31.86%) | 367(41.28%) |  |
| **Gender** |  |  | **0.036** |
| Male | 210 (66.25%) | 645 (72.55%) |  |
| Female | 106 (33.44%) | 241 (27.11%) |  |
| **Religion** |  |  |  |
| Christians (Overall) | 37 (11.67%) | 88 (9.90%) | ns |
| Christians (BAME) | 27 (8.52%) | 68 (7.65%) | ns |
| Hindu | 104 (32.81%) | 433 (48.71%) | **<0.0001** |
| Muslim | 134 (42.27%) | 253 (28.46%) | ref |
| **PHE guidance** |  |  | ns |
| Vulnerable | 79 (24.92%) | 224 (25.20%) |  |
| Healthy | 238 (75.08%) | 665 (74.80%) |  |
| **Ward area** |  |  | **0.0132** |
| Non-COVID-19 Only | 27 (8.52%) | 124 (13.95%) |  |
| COVID-19 | 290 (91.48%) | 765 (86.05%) |  |
| **Access to PPE** |  |  | **<0.0001** |
| Agree | 173 (54.57%) | 566 (63.67%) |  |
| Disagree | 116 (36.59%) | 199 (22.39%) |  |
| **Able to comply with Social Distancing** |  |  | **0.0203** |
| Most/all of the times | 96 (30.28%) | 334 (33.57%) |  |
| Some, few or none of the times | 221 (69.72%) | 555 (62.43%) |  |
| **Able to negotiate** |  |  | **0.0167** |
| None | 67 (21.14%) | 157 (17.66%) |  |
| Yes | 183 (57.73%) | 648 (72.89%) |  |
| **Reprimanded for PPE** |  |  | **<0.0001** |
| Yes | 123 (38.80%) | 237 (26.66%) |  |
| Rare/Never | 194 (61.20%) | 652 (73.34%) |  |
| **Redeployed to area that cares for** |  |  | ns |
| Non-COVID-19 only | 21 (6.63%) | 47 (5.29%) |  |
| COVID-19 only | 141 (44.48%) | 314 (35.32%) |  |

#### Multivariate analysis

**(a) Model I:** This model included Hindu and Muslim respondents only, thus excluding 282 respondents (125 Christians and 157 ‘others’ and with ‘no religious’ identity). In this model, none of the demographic variables were significant predictors of COVID-19. Out of the six variables determining occupational risk, inadequate PPE was an independent predictor for COVID-19 (OR 2.22 (95% CI 1.31 - 3.76, p = 0.003).

**(b) Model II**: This model compared Hindu (547), Muslim (387) and Christian-BAME respondents (95) (excluding 30 white respondents) (Table 5). In this model, none of the demographic variables were found to be significant predictors of COVID-19. Inadequate PPE remained the only independent predictor for self-reported COVID-19 (OR 2.29 (95% CI 1.22-4.33, p =0.01)).

**Table 5:**
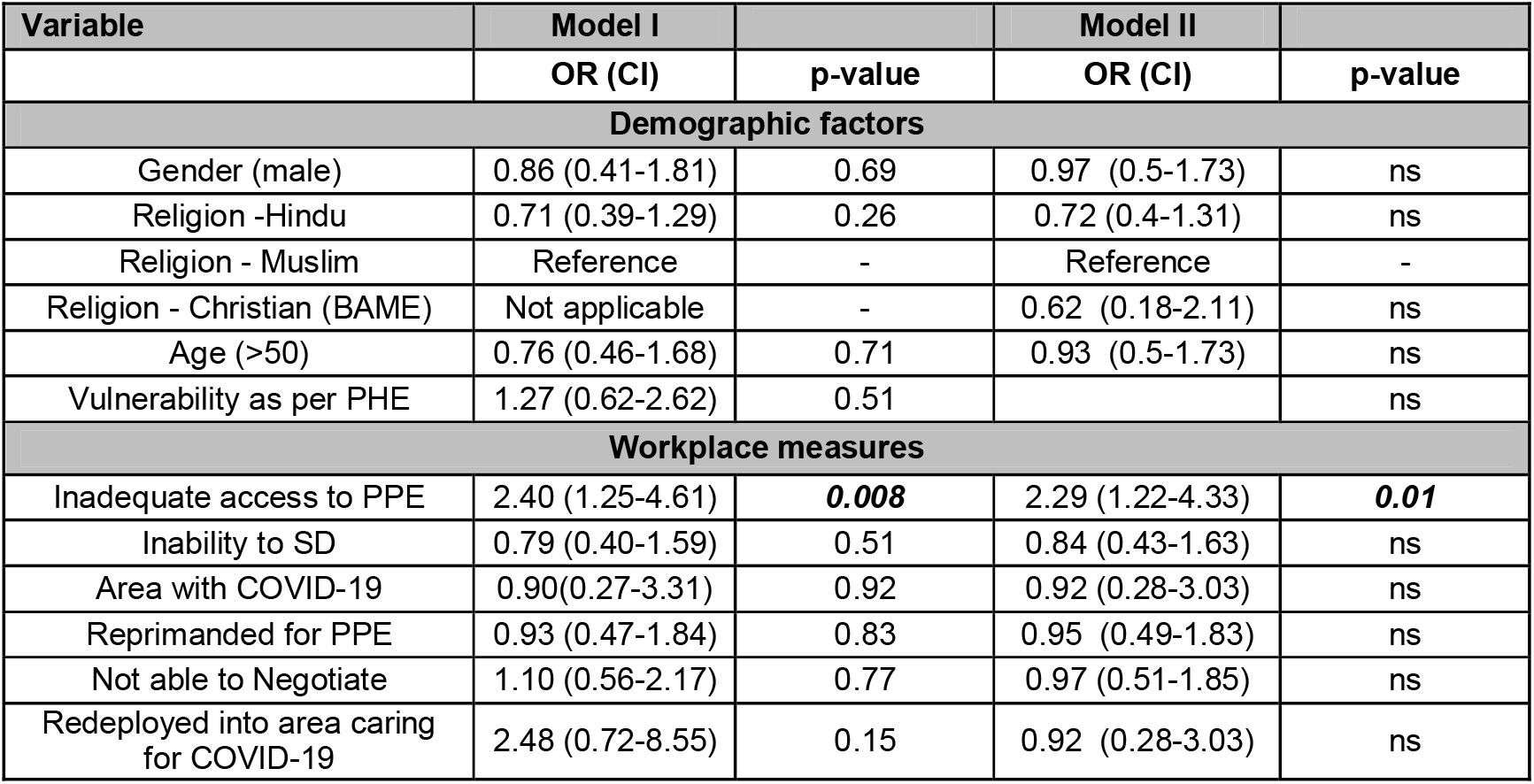
Binary Logistic regression analysis modelling for risk of COVID-19 Discussion.

| Variable | Model I |  | Model II |  |
| --- | --- | --- | --- | --- |
|  | OR (CI) | p-value | OR (CI) | p-value |
| **Demographic factors** |  |  |  |  |
| Gender (male) | 0.86 (0.41-1.81) | 0.69 | 0.97 (0.5-1.73) | ns |
| Religion -Hindu | 0.71 (0.39-1.29) | 0.26 | 0.72 (0.4-1.31) | ns |
| Religion - Muslim | Reference | - | Reference | - |
| Religion - Christian (BAME) | Not applicable | - | 0.62 (0.18-2.11) | ns |
| Age (>50) | 0.76 (0.46-1.68) | 0.71 | 0.93 (0.5-1.73) | ns |
| Vulnerability as per PHE | 1.27 (0.62-2.62) | 0.51 |  | ns |
| **Workplace measures** |  |  |  |  |
| Inadequate access to PPE | 2.40 (1.25-4.61) | **0.008** | 2.29 (1.22-4.33) | **0.01** |
| Inability to SD | 0.79 (0.40-1.59) | 0.51 | 0.84 (0.43-1.63) | ns |
| Area with COVID-19 | 0.90(0.27-3.31) | 0.92 | 0.92 (0.28-3.03) | ns |
| Reprimanded for PPE | 0.93 (0.47-1.84) | 0.83 | 0.95 (0.49-1.83) | ns |
| Not able to Negotiate | 1.10 (0.56-2.17) | 0.77 | 0.97 (0.51-1.85) | ns |
| Redeployed into area caring for COVID-19 | 2.48 (0.72-8.55) | 0.15 | 0.92 (0.28-3.03) | ns |

COVID-19 pandemic continues to be a major public health challenge. As far as we are aware, that this is the first survey that has studied gender and religion in the context of hospital doctors and risks of self-reported COVID-19. Hospital doctors are at an increased risk due to a higher exposure while caring for COVID-19 patients but also due to inconsistent access to appropriate PPE and compliance to SD at work (22,23,24). This is in addition to any applicable population-based risk factors (such as age, gender and comorbidities (4-9,12,24).

Many researchers have suggested risk assessment frameworks to minimise harm to those at highest risk (25,26) but these appear to be based on models of clinical risks and extrapolation of general population data. More recently, there has been a suggestion to include occupational factors in such a framework (27). We have previously reported data on HCWs including hospital doctors from the UK, demonstrating workplace measures and ethnicity were independent predictors of COVID-19 (22,23). The current study further explores additional characteristics such as gender and religion as risk factors COVID-19.

We found that women were more likely to report a diagnosis of COVID-19 but this was not found to be significant on multivariate analysis. Our study population included a higher proportion of women under 40 years of age (32% versus 17%), who were more likely to report a diagnosis of COVID-19. This may be representative of the demographics of the NHS frontline workforce (18,19). We found that gender and age were not independent predictors of COVID-19 in our study, in multivariate analysis. There is an excess risk of intensive care admissions and mortality from COVID-19 in men and those above 70 years (4). Some NHS trusts have already started risk stratification and are selectively redeploying BAME staff above 55 years, away from high risk areas (30,31).

This survey was open to all hospital doctors in the UK, however most responses received are likely from members of the two organisations representing doctors from Indian sub-continent heritage. Hence, it is not surprising that a significant majority of our respondents were from a BAME background and from three major religions practiced in the Indian sub-continent. Religious identity was not found to be statistically significant in determining risk, when adjusted for other factors in the multivariable analysis (Table 5).

Compliance with social distancing remains a challenge. Almost 2/3^rd^ of hospital doctors reported not being able to comply with social distancing and this was associated with increased risk of COVID-19. In the home, overcrowding and multi-generational households are also factors linked to higher exposure and hence increased risk to people from BAME background. Our survey in hospital doctors did not support this hypothesis. Data presented in the paper using a cut-off of five household numbers, but it was not significant even when analysing for a threshold of 2 and 3 (similar to average household numbers in UK (32). This could be because the socio-economic backgrounds of BAME hospital doctors are not comparable to the general population.

PPE is known to be one of the key measures ensuring safety of staff from occupational risk of COVID-19. There has been continued debate in the profession regarding the supply and timely delivery of appropriate PPE. We, and others have previously reported that many healthcare workers were not getting access to PPE as per PHE or WHO recommendations.[ref] In this survey, 61% hospital doctors reported appropriate access to PPE which is an improvement from 22% demonstrated previously (22,23). However, after adjusting for confounding variables, inadequate PPE remained an independent predictor with two-fold increased risk of COVID-19, in this cohort.

Lack of PPE may be associated with a degree of anxiety and stress for staff, in high risk clinical settings. In a previous survey by BMA, 64% of BAME staff felt pressured to work in settings with inadequate PPE (20). We found almost 30% hospital doctors reported being reprimanded for requesting PPE or risk avoidance measures (such as social distancing or redeployment in lower risk areas) and this was more commonly reported by Muslims. It would not be surprising that doctors facing discrimination are unlikely to raise concerns about inadequate workplace measures. The 2019 NHS staff survey and data from workforce race relations standards 2019 report (WRES) (18,19) indicates that overall 13% staff reported discrimination and another 31% reported facing bullying and undermining behaviour. The proportions were higher for BAME staff. Ethnicity was reported as the most common reason (eight times higher compared to the religious identity).

Our survey cohort is not directly comparable with the NHS staff survey as all our respondents were doctors and majority were male and BAME background, compared to 7.9% doctors 76% female and 20-40% from a BAME background). The fact that one-third of hospital doctors’ reported being reprimanded is deeply concerning. If hospital doctors (who have a more favourable educational and socio-economic background) report facing this degree of discrimination, it is likely that the experience may indeed be worse in other HCWs, more so from BAME backgrounds. This needs to be addressed by NHS organisations and staff support groups.

This study has a few limitations. A key comparator to workplace measures would have been between Caucasians and black ethnic respondents which had lower representation in our survey. General limitations to online surveys are also applicable to our survey.

## Conclusions

This survey contributes to the growing evidence of risk factors for COVID-19 amongst BAME doctors. Although the NHS has introduced risk assessment frameworks, these are based on demographics, and the scored on individual characteristics but not occupational or organisational influences. Access to PPE, although improved compared to results from April, still remains prevalent and inadequate access resulted in doubled the risk of COVID-19 for hospital doctors. Inability to comply with SD at work poses a similar challenge. Gender and religion did not contribute to additional risk, after adjusting to other variables in this study.

The unfortunate culture in the NHS, of being reprimanded or experiencing bullying and undermining contributes to an unsafe workplace for staff and where mistakes are more likely to lead to harm for patients. Hence, the focus needs to be on developing a culture of openness where the concerns can be raised safely and appropriate measures are taken to mitigate risks for staff and patients alike.

## Data Availability

yes available

## Author’s contribution and conflict of interest statements

Contributions are study design (SKD, GM, JSB & IC); data collection (All); analysis and interpreting the results (SKD & IC); writing the manuscript (SKD, SJ, MA, SG, IC); editing the manuscript and agreeing with final submission (All). The authors do not declare any conflict of interest.

## Acknowledgments

The authors wish to thank the staff at BAPIO Head Office, particularly Sneha Deshpande, members of BAPIO and APPNE and other organisations for supporting this survey.

